# Impact of Reproductive Factors and Hormone Replacement Therapy on Disease Severity in Women with Pulmonary Arterial Hypertension: Insights from the United States Pulmonary Hypertension Scientific Registry

**DOI:** 10.1101/2025.05.27.25328439

**Authors:** Jessica B. Badlam, Renee D. Stapleton, Peter W. Callas, David B. Badesch, Raymond L. Benza, Wendy K. Chung, Harrison W. Farber, Adaani E. Frost, Chang Yu, William C. Nichols, C. Gregory Elliott, Eric D. Austin

**Author notes:** ***Corresponding Author*** Jessica B. Badlam, MD, 89 Beaumont Ave, Given D213C, Burlington, VT 05405.

## Abstract

**Background:** Pulmonary arterial hypertension (PAH) is a devastating disease that affects women more often than men; recent U.S. cohorts demonstrate a female:male ratio of 3 to 4:1. Paradoxically, males have worse survival. The differential effects of sex hormones, particularly estrogen, on the pulmonary vasculature and right ventricle likely account for some of these differences. The role of female-specific risk factors, such as reproductive exposures, are poorly understood in the pathophysiology of PAH.

**Research Question:** To investigate the role of reproductive factors and exogenous estrogen exposures in PAH onset and severity in women enrolled in the United States Pulmonary Hypertension Scientific Registry (USPHSR).

**Study Design and Methods:** Using questionnaires from 390 women with PAH, enrolled in both the PAH Biobank and USPHSR, we conducted linear regression analyses to assess the association between patient reported reproductive variables and PAH disease severity variables, as well as REVEAL Lite 2.0 scores. We adjusted for potential confounders including age, race, BMI, and PAH sub-group (idiopathic, heritable, associated).

**Results:** Younger menopause age (< 40 years) associates with a lower cardiac index (CI) at diagnosis, even when controlling for use of hormone replacement therapy (HRT). There was a trend toward lower CI in women with menopause age of 41-50 years. Women who had ever used HRT were diagnosed with PAH an average of 13.4 years later and “ever use” of HRT associates with higher pulmonary vascular resistance and lower CI at diagnosis.

**Interpretation:** Premature menopause (age < 40 years) and ever use of HRT associate with worse hemodynamics, including lower CI, at diagnosis in women with PAH. Further investigations into reproductive history and estrogen exposures may offer an opportunity for more comprehensive risk factor screening and modification by physicians treating patients with PAH.

## Introduction

Pulmonary hypertension (PH) is elevated pressure in the pulmonary circulation (defined by mean PA pressure > 20mmHg) that can lead to right heart failure.^1^ Pulmonary arterial hypertension (PAH) is a progressive and fatal form of PH, with mean age at diagnosis of around 51 years.^2–4^ Female sex is a well-established risk factor for PAH, with recent US cohorts demonstrating an almost 4:1 female:male ratio.^3–5^ Despite this, males have worse survival.^5–7^ The exact reasons for these sex disparities in incidence and survival remain unclear.^8–10^ Studies have shown sex hormone levels (such as elevated estradiol [E2] and lower levels of dehydroepiandrosterone sulphate [DHEA-S]),^11^ altered metabolism,^12^ or signaling^13,14^ to be involved in the pathogenesis of the sexual dimorphism in PAH, with potentially disparate effects on the pulmonary vasculature and right ventricle. Few studies have investigated female sex-specific risk factors that alter endogenous exposure or the use of exogenous hormones and how they may confer additional risk in women.^15,16^ Identification of sex-specific risk factors may allow for earlier detection and/or individualized treatment of PAH.

There is increasing evidence that vascular disease in women, beyond PH, associates with sex-specific risk factors. Many of these factors reflect a woman’s lifetime exposure to estrogen, highlighting the potentially protective effects of E2. For example, atherosclerotic cardiovascular disease (CVD) risk in women is associated with reproductive factors such as age at menarche or menopause, infertility, presence of hypertensive disorders of pregnancy, type and duration of exogenous estrogen therapy, and absence of breastfeeding.^17^ The menopausal transition in women is characterized by a marked decline in levels of E2 and other sex hormones and is associated with accelerated CVD risk.^18^ These are unique factors in women and not yet fully investigated for associations and mechanistic links to the female predominance of PAH.

The United States Pulmonary Hypertension Scientific Registry (USPHSR) is a unique multi-center, longitudinal study, linked with the NHLBI sponsored National Biological Sample and Data Repository for PAH (PAH Biobank), that enrolled 499 patients diagnosed with PAH between 2012 and 2018. This registry contains demographic, physiologic, reproductive history, hormone exposure, and genomic data in the modern therapeutic era and was established to explore the potential genetic and environmental risk factors underlying the female predominance in PAH.^19^ In the USPHSR, we found that idiopathic PAH (IPAH) patients had significantly higher rates of miscarriage and oral contraceptive (OCP) use compared to those with other forms of PAH, suggesting a role for both reproductive factors and exogenous hormone exposure in the pathophysiology of IPAH.^3^ Additionally, post-menopausal status and/or hormone replacement therapy (HRT) use may affect PAH disease severity.^15^ A cohort study using the UK Biobank found that premature menopause may be an independent risk factor for PH in women.^21^ Given that not everyone with a known risk factor for the development of PAH develops disease, it is critical to understand how these reproductive factors contribute to the pathogenesis or progression of disease.

We sought to understand the relationship between sex-specific factors and the development and severity of PAH. We investigated female specific risk factors, including reproductive factors and exogenous hormone use, in women in the USPHSR to determine the association between these exposures and PAH severity.

## Materials and Methods

### STUDY SUBJECTS

USPHSR interviewers administered an expanded version of the previously validated and extensively published Nurses’ Health Study (NHS) I and II questionnaire^21^ to all 499 adult (≥ 18 years old) participants (392 women, 107 men). These questionnaires were designed to address a participant’s reproductive history via questions related to menstruation, menopause, pregnancy, breast-feeding, and infertility. This information is available on 390 of the 392 female patients with PAH enrolled in USPHSR: 133 with IPAH, 57 with heritable PAH (HPAH), and 200 with associated PAH (APAH).^3^ At the time of USPHSR enrollment, additional data were obtained from a patient interview, medical record review, and data stored at the PAH Biobank. Baseline demographic elements were collected and entered into electronic case report forms. N-terminal pro B-type natriuretic peptide (NTproBNP) levels were measured on PAH Biobank biospecimens by the Johns Hopkins Pediatric Proteome Center.

### METHODS/ANALYSIS

We conducted linear regression analyses to assess the association between age and disease severity variables at diagnosis and reproductive variables that reflect lifetime endogenous estrogen exposure. Age at diagnosis and PAH disease severity variables at diagnosis were obtained from the PAH Biobank including WHO functional class, six minute walk distance, and right heart catheterization hemodynamics (right atrial pressure [RAP], mean pulmonary artery pressure [mPAP], pulmonary vascular resistance [PVR], cardiac index [CI]). The reproductive factors were the predictors and included number of pregnancies, ever having miscarriage, ever having premature birth, ever breastfeeding, menarche age, menopause age, surgical menopause/bilateral oophorectomy, and use of fertility medications. We then conducted linear regression analyses to assess the association between age/PAH disease severity variables at diagnosis and “ever use” of OCP or HRT. We adjusted for potential confounders including age, race/ethnicity, body mass index (BMI), other reproductive factors, and PAH sub-group (idiopathic, heritable, or associated). A sample size of 390 subjects provided 84% power to detect a correlation coefficient of 0.15 (with a two-sided type I error rate of 0.05). In addition, this sample size was large enough to ensure that we exceeded the commonly used criterion of 10 observations per covariate in a multivariable linear model.

We also investigated associations between duration of OCP or HRT use and PAH disease severity/mortality risk at diagnosis determined by the validated REVEAL Lite 2.0 risk calculator.^14^ The REVEAL Lite 2.0 calculator requires a minimum of 3 of the following variables to generate a score where 2 are the most predictive variables (*italicized*); *B-type natriuretic peptide (BNP)* or *NTproBNP, six minute walk test, NYHA/WHO functional class*, systolic blood pressure, heart rate, and renal function. Higher scores indicate a greater risk of 1-year mortality from PAH. After generating the scores from available data, we then used multivariable linear regression to investigate the relationship of duration of OCP or HRT use with age at diagnosis or REVEAL Lite 2.0 risk score. We adjusted for potential confounders including age, race, BMI, PAH sub-group, and menopausal status.

All statistical analyses were performed using SAS version 9.4, and p<0.05 was considered statistically significant. (Cary, NC: SAS Institute Inc.)

## Results

### Reproductive Factors

For the 390 women enrolled in USPHSR who completed the questionnaire, the mean age at PAH diagnosis was 52 years and mean age at enrollment was 56 years. We previously described the reported reproductive factors and endogenous sex hormone exposures in female USPHSR subjects according to their PAH classification.^3^ **Table 1** depicts the association between age and disease severity variables at diagnosis and reproductive variables. Overall, we found that two reproductive variables, ever having miscarriage and menopause age, associated with hemodynamics at the time of PAH diagnosis.

**Table 1.** Association between age and disease severity variables at PAH diagnosis and reproductive variables in female USPHSR participants.

| Table 1. |  | Age at Diagnosis (y)<br>n = 389 |  |  | Functional Class (N)<br>n = 317 |  |  | Six Minute Walk Distance<br>(m) n = 297 |  |  | Right Atrial Pressure<br>(mmHg)<br>n = 382 |  |  |
| --- | --- | --- | --- | --- | --- | --- | --- | --- | --- | --- | --- | --- | --- |
| | | Mean<sup>+</sup><br>(SD) | $\beta$<br>(SE) | P<br>value | Mean<br>(SD) | $\beta$ (SE) | P<br>value | Mean<br>(SD) | $\beta$ (SE) | P<br>value | Mean<br>(SD) | $\beta$ (SE) | P<br>value |
|  |  | 52.0<br>(15.3) |  |  | 2.73<br>(0.69) |  |  | 331<br>(109) |  |  | 9.2<br>(5.8) |  |  |
| Reproductive Variable | Answer |  |  |  |  |  |  |  |  |  |  |  |  |
| Number of Pregnancies | N |  | -0.1<br>(0.1) | 0.24 |  | 0.00<br>(0.03) | 0.91 |  | 7 (5) | 0.18 |  | -0.3<br>(0.2) | 0.28 |
| Ever had miscarriage | Yes | 53.4<br>(14.6) | 0.0<br>(0.2) | 0.97 | 2.70<br>(0.69) | -0.10<br>(0.11) | 0.36 | 327<br>(107) | -10 (17) | 0.56 | 9.2<br>(6.1) | -0.1<br>(0.8) | 0.88 |
|  | No | 51.3<br>(15.7) |  |  | 2.74<br>(0.69) |  |  | 332<br>(111) |  |  | 9.3<br>(5.7) |  |  |
| Ever had premature birth | Yes | 51.0<br>(14.7) | 0.3<br>(0.2) | 0.19 | 2.63<br>(0.75) | -0.16<br>(0.11) | 0.15 | 345<br>(122) | -4 (19) | 0.81 | 9.2<br>(5.2) | 0.2 (0.9) | 0.82 |
|  | No | 52.2<br>(15.5) |  |  | 2.75<br>(0.67) |  |  | 328<br>(106) |  |  | 9.3<br>(5.9) |  |  |
| Ever breastfed | Yes | 51.0<br>(14.4) | 0.1<br>(0.2) | 0.59 | 2.72<br>(0.70) | -0.03<br>(0.09) | 0.78 | 346<br>(118) | 8 (15) | 0.57 | 9.0<br>(6.0) | -0.2<br>(0.7) | 0.82 |
|  | No | 52.6<br>(15.9) |  |  | 2.73<br>(0.68) |  |  | 320<br>(102) |  |  | 9.4<br>(5.6) |  |  |
| Menarche Age | N |  | 0.0<br>(0.1) | 0.49 |  | 0.01<br>(0.03) | 0.75 |  | 4 (4) | 0.38 |  | -0.2<br>(0.7) | 0.82 |
| Menopause Age | NA | 42.5<br>(14.1) | 0.0<br>(0.3) | 0.88 | 2.78<br>(0.70) | -0.12<br>(0.15) | 0.42 | 354<br>(105) | 11 (25) | 0.67 | 9.3<br>(5.5) | -0.5<br>(1.1) | 0.66 |
|  | Unknown | 63.2<br>(9.2) | 0.1<br>(0.4) | 0.80 | 2.71<br>(0.64) | 0.07<br>(0.17) | 0.68 | 304<br>(114) | -23 (30) | 0.44 | 9.5<br>(6.0) | 1.6 (1.4) | 0.24 |
|  | < 40 | 59.7<br>(11.1) | 0.0<br>(0.5) | 0.97 | 2.50<br>(0.63) | -0.19<br>(0.22) | 0.37 | 275<br>(112) | -10 (36) | 0.79 | 8.6<br>(4.7) | -0.3<br>(1.7) | 0.87 |
|  | 40-44 | 61.0<br>(10.7) | 0.0<br>(0.4) | 0.92 | 2.64<br>(0.75) | 0.04<br>(0.21) | 0.86 | 284<br>(82) | -22 (32) | 0.49 | 12.7<br>(9.7) | 4.9 (1.5) | <0.001 |
|  | 45-49 | 61.5<br>(9.4) | -0.5<br>(0.3) | 0.10 | 2.71<br>(0.75) | -0.01<br>(0.15) | 0.96 | 315<br>(102) | 15 (26) | 0.57 | 9.2<br>(5.7) | 0.7 (1.2) | 0.54 |
|  | ≥ 50 | 63.5<br>(8.3) |  |  | 2.71<br>(0.66) | Referent |  | 317<br>(115) | Referent |  | 7.8<br>(4.7) | Referent |  |
| Bilateral oophorectomy | Yes | 59.7<br>(10.9) | -0.3<br>(0.3) | 0.30 | 2.75<br>(0.64) | 0.10<br>(0.12) | 0.41 | 292<br>(96) | -18 (19) | 0.33 | 9.7<br>(5.9) | 0.9 (0.9) | 0.33 |
|  | No | 50.4<br>(15.6) |  |  | 2.73<br>(0.70) |  |  | 339<br>(110) |  |  | 9.1<br>(5.8) |  |  |
| Use of Fertility Medications | Yes | 46.3<br>(11.9) | -0.3<br>(0.4) | 0.49 | 2.65<br>(0.61) | -0.06<br>(0.18) | 0.73 | 416<br>(78) | 66 (28) | 0.02 | 9.0<br>(4.8) | 0.5 (1.5) | 0.74 |
|  | No | 52.2<br>(15.4) |  |  | 2.73<br>(0.69) |  |  | 326<br>(109) |  |  | 9.2<br>(5.8) |  |  |

| Table 1 continued. |  | Mean Pulmonary Artery Pressure (mmHg)<br>n = 389 |  |  | Pulmonary Vascular Resistance (Wood units)<br>n = 374 |  |  | Cardiac Index (L/min/m<sup>2</sup>)<br>n = 376 |  |  |
| --- | --- | --- | --- | --- | --- | --- | --- | --- | --- | --- |
|  |  | Mean* (SD) | β (SE) | P value | Mean (SD) | β (SE) | P value | Mean (SD) | β (SE) | P value |
|  |  | 49.0 (13.1) |  |  | 10.5 (5.9) |  |  | 2.35 (0.8) |  |  |
| **Reproductive Variables** | **Answer** |  |  |  |  |  |  |  |  |  |
| **Number of Pregnancies** | N |  | 0.1 (0.5) | 0.84 |  | 0.3 (0.2) | 0.19 |  | -0.04 (0.03) | 0.25 |
| **Ever had miscarriage** | Yes | 47.3 (12.5) | -2.7 (1.8) | 0.14 | 9.8 (5.7) | -1.6 (0.8) | **0.05** | 2.41 (0.75) | 0.19 (0.11) | 0.10 |
|  | No | 49.8 (13.2) |  |  | 10.8 (6.0) |  |  | 2.34 (0.82) |  |  |
| **Ever had premature birth** | Yes | 50.4 (13.6) | 1.9 (1.9) | 0.33 | 10.7 (6.3) | 0.3 (0.9) | 0.75 | 2.35 (0.69) | 0.03 (0.12) | 0.83 |
|  | No | 48.7 (12.9) |  |  | 10.5 (5.8) |  |  | 2.36 (0.83) |  |  |
| **Ever breastfed** | Yes | 48.6 (13.2) | -1.6 (1.5) | 0.30 | 10.8 (6.4) | 0.1 (0.7) | 0.89 | 2.31 (0.76) | -0.07 (0.10) | 0.46 |
|  | No | 49.3 (13.0) |  |  | 10.3 (5.6) |  |  | 2.39 (0.83) |  |  |
| **Menarche Age** | N |  | -0.3 (0.5) | 0.51 |  | -0.3 (0.2) | 0.11 |  | 0.02 (0.03) | 0.55 |
| **Menopause Age** | NA | 51.4 (13.1) | -0.4 (2.5) | 0.87 | 9.8 (6.3) | -0.7 (1.1) | 0.55 | 2.31 (0.72) | -0.15 (0.15) | 0.33 |
|  | Unknown | 49.2 (13.9) | -0.3 (3.0) | 0.92 | 10.4 (5.3) | -0.4 (1.4) | 0.78 | 2.47 (0.92) | 0.05 (0.19) | 0.78 |
|  | < 40 | 46.9 (9.7) | 0.7 (3.8) | 0.85 | 10.4 (5.9) | 1.4 (1.7) | 0.41 | 2.06 (0.65) | -0.57 (0.24) | **0.02** |
|  | 40-44 | 45.2 (12.1) | -1.2 (3.3) | 0.71 | 9.1 (4.5) | -0.5 (1.5) | 0.76 | 2.32 (0.94) | -0.34 (0.20) | 0.10 |
|  | 45-49 | 47.9 (12.4) | 2.1 (2.7) | 0.44 | 10.4 (5.7) | 1.4 (1.2) | 0.24 | 2.33 (0.81) | -0.29 (0.17) | 0.09 |
|  | ≥ 50 | 46.1 (14.0) | Referent |  | 9.0 (6.2) | Referent |  | 2.59 (0.91) | Referent |  |
| **Bilateral oophorectomy** | Yes | 45.0 (10.6) | -2.7 (2.0) | 0.18 | 9.5 (5.5) | 0.2 (0.9) | 0.87 | 2.29 (0.78) | -0.05 (0.12) | 0.69 |
|  | No | 49.9 (13.4) |  |  | 10.8 (6.0) |  |  | 2.37 (0.81) |  |  |
| **Use of Fertility Medications** | Yes | 46.6 (11.1) | -3.9 (3.4) | 0.25 | 9.1 (5.4) | -2.0 (1.5) | 0.19 | 2.37 (0.70) | 0.07 (0.21) | 0.74 |
|  | No | 49.2 (13.2) |  |  | 10.6 (5.9) |  |  | 2.35 (0.81) |  |  |

Abbreviations: SD – Standard Deviation; SE- Standard Error; NA – Not applicable, participants pre-menopausal; m -metres; y – years; mmHg – millimetres of mercury; L – litres; min – minute
\*Means are for unadjusted values; β (SE) represent adjusted variables for age, race, body mass index, PAH subgroup, and other reproductive variables

Ever having a miscarriage associated with lower PVR at diagnosis (mean -1.6 Wood units, p=0.05). There was no association of prior miscarriage with other hemodynamics such as mPAP or CI. We also found that menopausal status, particularly premature menopause at <40 years, associated with lower CI at diagnosis (mean -0.57 L/min/m^2^, p=0.02) when compared to a menopause age of ≥ 50 years, and remained significant even when controlling for use of HRT. There was also a trend toward lower CI (−0.3 L/min/m^2^) in those females with a menopause age of 40-49 years old, but this was not statistically significant. There was no association with menopause age and other hemodynamics such as mPAP or PVR. Additionally, we found no association of age at diagnosis or other disease severity variables with additional reproductive factors (number of pregnancies, ever had premature birth or breastfed, menarche age, or surgical menopause).

### Exogenous Estrogen Use

We previously described the reported exogenous sex hormone exposures in female USPHSR participants according to their PAH classification.^3^ **Table 2** depicts the association between age and disease severity variables at diagnosis and ever use of OCP or HRT. Compared with women who reported no prior use of HRT, those women who had ever used HRT were diagnosed with PAH an average of 13.4 years later. Additionally, self-report of ever use of HRT was associated with worse hemodynamics at PAH diagnosis, including a higher PVR by 1.6 Wood units (p=0.05) and lower CI at -0.28 L/min/m^2^ (p=0.01). Similar trends occurred when we limited the cohort to females who also self-reported being post-menopausal at the time of query, but this was not statistically significant.

**Table 2.** Association between age and disease severity variables at PAH diagnosis and ever use of exogenous estrogen in female USPHSR participants.

| Table 2. |  | Age at Diagnosis (y)<br>n = 386 |  |  | Functional Class (N)<br>n = 315 |  |  | Six Minute Walk<br>Distance (m)<br>n = 296 |  |  | Right Atrial Pressure<br>(mmHg)<br>n = 379 |  |  |
| --- | --- | --- | --- | --- | --- | --- | --- | --- | --- | --- | --- | --- | --- |
| | | Mean*<br>(SD) | $\beta$<br>(SE) | P<br>value | Mean<br>(SD) | $\beta$ (SE) | P<br>value | Mean<br>(SD) | $\beta$<br>(SE) | P<br>value | Mean<br>(SD) | $\beta$<br>(SE) | P<br>value |
|  |  | 52.0<br>(15.3) |  |  | 2.73<br>(0.69) |  |  | 331<br>(109) |  |  | 9.2<br>(5.8) |  |  |
|  | **Answer** |  |  |  |  |  |  |  |  |  |  |  |  |
| **Ever use of OCP** | No | 55.4<br>(17.7) | 0.1<br>(0.2) | 0.50 | 2.74<br>(0.67) | -0.05<br>(0.09) | 0.58 | 327<br>(90) | -10<br>(14) | 0.48 | 8.7<br>(5.5) | 0.5<br>(0.7) | 0.50 |
|  | Yes | 51.0<br>(14.3) |  |  | 2.72<br>(0.69) |  |  | 332<br>(114) |  |  | 9.4<br>(5.9) |  |  |
| **Ever use of HRT** | No | 49.0<br>(14.9) | 13.4<br>(1.8) | **<0.001** | 2.72<br>(0.70) | 0.11<br>(0.10) | 0.28 | 337<br>(113) | 0<br>(17) | 0.98 | 9.3<br>(5.7) | 0.5<br>(0.8) | 0.52 |
|  | Yes | 63.4<br>(10.5) |  |  | 2.69<br>(0.63) |  |  | 312<br>(96) |  |  | 8.8<br>(5.7) |  |  |

| Table 2 continued. |  | Mean Pulmonary Artery<br>Pressure (mmHg)<br>n = 386 |  |  | Pulmonary Vascular<br>Resistance (Wood units)<br>n = 372 |  |  | Cardiac Index (L/min/m<sup>2</sup>)<br>n = 374 |  |  |
| --- | --- | --- | --- | --- | --- | --- | --- | --- | --- | --- |
| | | Mean<br>(SD) | $\beta$ (SE) | P<br>value | Mean (SD) | $\beta$ (SE) | P<br>value | Mean<br>(SD) | $\beta$ (SE) | P<br>value |
|  |  | 49.0<br>(13.1) |  |  | 10.5 (5.9) |  |  | 2.35 (0.8) |  |  |
|  | **Answer** |  |  |  |  |  |  |  |  |  |
| **Ever use of OCP** | No | 47.5<br>(11.9) | 0.5<br>(1.6) | 0.73 | 10.3 (6.3) | -0.3<br>(0.7) | 0.69 | 2.46<br>(0.92) | -0.09<br>(0.10) | 0.37 |
|  | Yes | 49.6<br>(13.4) |  |  | 10.6 (5.8) |  |  | 2.32<br>(0.76) |  |  |
| **Ever use of HRT** | No | 49.7<br>(13.3) | 0.4<br>(1.8) | 0.82 | 10.6 (5.9) | 1.6<br>(0.8) | **0.05** | 2.39<br>(0.83) | -0.28<br>(0.11) | **0.01** |
|  | Yes | 46.6<br>(12.5) |  |  | 10.6 (6.2) |  |  | 2.21<br>(0.67) |  |  |

Abbreviations: OCP – Oral Contraceptive Pill; HRT – Hormone Replacement Therapy; SD – Standard Deviation; SE- Standard Error; m -metres; y – years; mmHg – millimetres of mercury; L – litres; min – minute
\*Means are for unadjusted values; $\beta$ (SE) represent adjusted variables for age, race, body mass index, PAH subgroup

We were able to calculate REVEAL Lite 2.0 scores for 349 of the 390 women in USPHSR based on available data. The average score was 6.4 +/-2.2. There were no associations found between duration of exogenous estrogen use (OCP or HRT) and age at diagnosis or REVEAL Lite 2.0 score (**Table 3**), including for duration of HRT use when we only included females who also self-reported being post-menopausal. We also conducted similar analyses of reproductive factors (number of pregnancies, ever had miscarriage/premature birth/breastfed, menarche age, menopause age, or surgical menopause) and REVEAL Lite 2.0 scores and found no significant associations.

**Table 3.** Association between age and REVEAL Lite 2.0 score at PAH diagnosis and duration of exogenous estrogen use in female USPHSR participants.

| Table 3. |  | Age at Diagnosis (y)<br>n = 378 |  |  | REVEAL Lite 2.0 Score (N)<br>n = 339 |  |  |
| --- | --- | --- | --- | --- | --- | --- | --- |
| | | Mean (SD) | $\beta$ (SE) | P value | Mean (SD) | $\beta$ (SE) | P value |
|  |  | 52.0 (15.3) |  |  | 6.4 (2.2) |  |  |
|  | **Answer** |  |  |  |  |  |  |
| **Duration of OCP use** | Never | 55.4 (17.7) | Referent |  | 6.7 (2.4) | Referent |  |
|  | <1 year | 53.2 (14.2) | 0.4 (0.3) | 0.15 | 7.0 (2.1) | 0.5 (0.5) | 0.27 |
|  | 1-5 years | 53.0 (15.0) | 0.1 (0.2) | 0.64 | 6.2 (2.0) | -0.3 (0.3) | 0.33 |
|  | 5-10 years | 45.6 (14.5) | -0.2 (0.3) | 0.46 | 6.2 (2.3) | 0.0 (0.4) | 0.98 |
|  | 10-15 years | 51.6 (13.1) | 0.2 (0.3) | 0.61 | 6.6 (2.4) | 0.3 (0.4) | 0.49 |
|  | 15-20 years | 50.4 (11.5) | 0.5 (0.5) | 0.25 | 5.7 (2.7) | -0.7 (0.7) | 0.28 |
|  | >20 years | 49.3 (10.2) | 0.6 (0.5) | 0.18 | 6.4 (2.0) | -0.1 (0.7) | 0.88 |
| **Duration of HRT use** | Never | 49.0 (14.9) | Referent |  | 6.3 (2.3) | Referent |  |
|  | <1 year | 58.5 (10.6) | 0.4 (0.6) | 0.52 | 5.7 (0.9) | -0.9 (0.8) | 0.23 |
|  | 1-5 years | 66.0 (9.4) | -0.1 (0.3) | 0.66 | 6.8 (2.0) | -0.1 (0.4) | 0.89 |
|  | 5-10 years | 59.6 (12.9) | -0.4 (0.5) | 0.45 | 7.4 (2.3) | 0.9 (0.7) | 0.20 |
|  | 10-15 years | 65.9 (6.8) | -0.5 (0.7) | 0.52 | 7.4 (1.5) | 0.4 (1.0) | 0.68 |
|  | 15-20 years | 64.5 (3.5) | -0.5 (0.6) | 0.45 | 5.2 (1.9) | -1.7 (1.0) | 0.08 |
|  | >20 years | 66.5 (12.1) | 0.5 (0.7) | 0.44 | 6.3 (2.1) | -0.3 (0.9) | 0.74 |

Abbreviations: OCP – Oral Contraceptive Pill; HRT – Hormone Replacement Therapy; SD – Standard Deviation; SE – Standard Error; y – years
\*Means are for unadjusted values; $\beta$ (SE) represent adjusted variables for age, race, body mass index, PAH subgroup

## Discussion

Female sex is one of the most well-established risk factors for PAH, but females have better survival than males.^8^ There are many reasons why this may be the case, including the effects of estrogen synthesis, metabolism, and signaling on the pulmonary vasculature and right heart.^26^ We sought to evaluate how a woman’s lifetime exposure to both endogenous and exogenous estrogen affects PAH age of onset and severity at time of diagnosis. We found that in female patients in the USPHSR, a self-reported history of ever having a miscarriage and premature menopause both associated with significant differences in hemodynamics at diagnosis of PAH. We found that the ever use of HRT associates with a later age at diagnosis but also significantly associates with worse hemodynamics at diagnosis. There was no association between non-invasive REVEAL Lite 2.0 risk scores and reproductive factors or use of exogenous estrogen.

Pregnancy, including those that end in miscarriage or stillbirth, reduce a woman’s total duration of endogenous estrogen exposure. Miscarriage is a common outcome of pregnancy, affecting upwards of 10-20% of all recognized pregnancies, and this rate increases with age.^27^ Common etiologies of pregnancy loss include chromosomal abnormalities, maternal anatomic abnormalities, and trauma, but there are various maternal morbidities such as obesity, diabetes, and thyroid disease that associate with pregnancy loss as well.^30^ We previously demonstrated that females with IPAH had higher rates of miscarriage compared to other etiologies of PAH even with similar parity.^3^ Despite no association of parity with PAH disease severity in this current study, we did find that ever having miscarriage associates with lower PVR at diagnosis. The implications of these findings are not entirely clear but raise the question of how endogenous estrogen exposure may influence vascular disease manifestations in women prior to the diagnosis of PAH.^30^

Menopause is defined as the permanent cessation of menses, resulting from the depletion of potentially functional primordial follicles in the ovaries and relative estrogen deficiency.^26^ Premature menopause or primary ovarian insufficiency is the development of amenorrhea, symptoms of estrogen deficiency, and gonadotropin levels in the menopausal range before age 40 years. The cause is frequently unknown but may be the result of genetic abnormalities, autoimmune disease, surgical removal of the ovaries, or treatment such as chemotherapy that may damage ovarian tissue. Notably, premature menopause appears to place females at a variety of increased risk including osteoporosis and fractures^27^, CVD^28^ including pulmonary hypertension^20^, and premature death (total mortality and mortality from ischemic heart disease).^29,30^

Our study demonstrated a history of premature menopause associates with a lower cardiac index at PAH diagnosis, indicating worse right ventricular (RV) function. There was also a trend toward lower CI at diagnosis in early menopause (< 45 years). RV function is a critical factor in the sexually dimorphic outcomes in PAH.^7^ These findings are consistent with previous studies demonstrating baseline higher CI in females than in males with idiopathic PAH^7,31^ and the plethora of pre-clinical studies supporting that sex differences in mortality are due in part to the beneficial effects of E2 on RV function.^8^ Premature menopause appears to be a risk factor for the development of PH^20^ and potentially more severe PAH in females and may be an opportunity for earlier identification of or screening for pulmonary vascular disease in females.

It must also be noted that the average age of menopause in females is around ∼52 years.^26^ The average age of diagnosis of PAH, in both males and females, is approximately 52 years in more recent US cohorts, including REVEAL^32^ and USPHSR. The similar timing of these events and sex hormone fluctuations during the perimenopause transition in females is unlikely to be coincidental. Alterations in endogenous estrogen synthesis in this time period can affect both perception of physical function and true physical performance.^33,34^ This may be, in part, the result of altered vascular endothelial and RV myocyte function.^35^ All of these factors likely contribute to the development of symptoms and PAH disease presentation during the perimenopause transition.

We found that in the USPHSR female population, ever use of HRT associated with more severe hemodynamics at diagnosis including higher PVR and lower CI. We did not find any association with reported duration of HRT use and REVEAL Lite 2.0 scores. This is in contrast to prior findings in healthy post-menopausal women in the MESA cohort that had better RV function with higher E2 levels and use of HRT^36^ and recently presented data from the PVDOmics study that women with PH including PAH had lower mPAP and PVR and higher RV ejection fraction with past HRT exposure.^37^ The dose, duration, and timing of HRT used in each of these populations is unknown and certainly may account for the incongruent results. These data may prove to be essential as the risks and benefits of HRT use in CVD have been evaluated extensively over the past three decades resulting in “The Timing Hypothesis” that the effects of HRT depend on the timing of initiation of HRT in relation to menopause.^38^ The effect of exogenous estrogen on the pulmonary vasculature and right ventricular function varies in pre-clinical and human studies and is likely affected by multiple other factors including estrogen metabolism, other sex hormones, and non-hormonal factors such as sex chromosomes, epigenetic modifications and inflammation/immune cell regulation.^10^ Better understanding of these complicated interactions will certainly add to our understanding of the risks and benefits of HRT in females with PAH.

This study provides novel insights on endogenous and exogenous estrogen exposures in females with PAH but has limitations. We acknowledge that this is an observational study and cannot infer causality of these exposures. Many unmeasured factors (such as socioeconomic status, geographical trends in prescribing, etc) contribute to reproductive outcomes and are unaccounted for in this study. There are known limitations to the questionnaire used to assess reproductive history, such as type/dose of OCP or HRT used and timing of exposure, as there was a need to balance the depth and number of questions with the likelihood of completion of the questionnaire. Given the lag time between diagnosis and enrolment in USPHSR, ∼4 years, the timing of these exposures in relation to diagnostic testing is unknown and introduces survivor bias as most were prevalent cases. Patient responses to the questionnaire are also limited by self-reporting and recall bias. As there is no standard way to determine lifetime endogenous estrogen exposure, these factors were assessed independently in this study; future studies could consider composite exposure risk.

Overall, we found that some reproductive factors related to lifetime estrogen exposure in females with PAH associate with worse hemodynamics at diagnosis. Premature menopause (age < 40 years) associated with worse RV function in women with PAH, including lower CI. Ever use of HRT also associated with more severe hemodynamics at diagnosis, including higher PVR and lower CI. Further investigations into reproductive factors and estrogen exposures may offer a unique opportunity for more comprehensive risk factor screening and/or modification in in this deadly disease. The impact of the broader sex hormone and metabolite milieu in PAH cannot be ignored and we look forward to seeing how this influences future treatment options for patients with PAH.

## Data Availability

All data produced in the present study are available upon reasonable request to the authors.

